# Microbiome compositions and fecal metabolite concentrations predict post-operative infection in liver transplant recipients

**DOI:** 10.1101/2023.02.17.23286090

**Authors:** Christopher J Lehmann, Nicholas P Dylla, Matthew Odenwald, Ravi Nayak, Maryam Khalid, Jaye Boissiere, Jackelyn Cantoral, Emerald Adler, Matthew R Stutz, Mark Dela Cruz, Angelica Moran, Huaiying Lin, Anitha Sundararajan, Ashley M. Sidebottom, Eric G Pamer, Andrew Aronsohn, John Fung, Talia B Baker, Aalok Kacha

**Author notes:** Christopher J Lehmann, MD^1^, 5841 South Maryland Ave., MC 5065, Chicago, IL 60637. Eric G Pamer^1,2^. Aalok Kacha^9^.

## Abstract

**Background:** Liver transplantation (LT) is associated with postoperative infections caused by antibiotic-resistant bacterial pathogens that reside in the intestine. An intact intestinal microbiome suppresses expansion of enteric pathogens, however patients with severe liver disease often have reduced microbiome diversity and increased density of antibiotic-resistant *Enterococcus* and *Enterobacterales* species. (1–4) Experimental models have demonstrated that metabolites produced by the intestinal microbiome, including short chain fatty acids (SCFAs), secondary bile acids and indole compounds, enhance host epithelial and immune defenses against enteric pathogens.(5–10) Microbiome derived metabolites likely contribute to resistance against infectious diseases in LT patients, however, this remains uninvestigated.

**Methods:** We prospectively enrolled 107 liver transplant candidates and determined peri-transplant fecal microbiome compositions including relative and absolute fecal metabolite concentrations.

**Results:** Fecal microbiomes in LT recipients ranged from highly diverse to complete loss of diversity resulting in expansion of *Enterococcus* and/or *Enterobacterales* species that were associated with postoperative infection. Gas chromatographic (GC-) and liquid chromatographic (LC-) Mass spectrometric analyses revealed decreased concentrations of SCFAs, secondary bile acids, and indole compounds in fecal samples with low microbiome diversity and associated expansion of *Enterococcus* and *Enterobacterales* populations.

**Conclusion:** Fecal metabolite abundances accurately predicted LT patients with reduced microbial diversity and those who developed postoperative infection.

## INTRODUCTION

Liver transplantation (LT) is the definitive treatment for end-stage liver disease. Despite advances in antibiotic prophylaxis, patient selection, donor screening, organ preservation, and surgical technique, infection remains a leading cause of morbidity and mortality. (11–13) *Enterococcus* species and members of the order *Enterobacterales*, such as *Escherichia coli* and *Klebsiella pneumonia*, remain among the most common causes of infection in the post-transplant period. The clinical impact of these infections is compounded by rising rates of antibiotic resistance, with vancomycin resistant *Enterococcus faecium* (VRE) and multiple drug resistant *Enterobacterales* making outsized contributions to morbidity and mortality. (12, 13)

*Enterococcus* and *Enterobacterales* species are normal inhabitants of the human gut microbiota. (14, 15) In healthy individuals, these facultative anaerobes compose a small fraction of the microbiota but are capable of markedly expanding following loss of microbiota diversity. (16, 17) Pathogen expansion in the gut increases the risk of systemic infection in a variety of disease states. (18, 19)How the microbiome suppresses *Enterococcus* and *Enterobacterales* expansion in the gut and prevents systemic infections is an active area of investigation with increasing evidence supporting the role of microbiota-derived metabolites, especially SCFAs, secondary bile acids, and indole compounds in bacterial growth inhibition and enhancement of host immune and epithelial barrier defenses. (20–24)

The intestinal microbiota is impacted by liver disease and gut microbial populations produce metabolites and other products that circulate via the portal vein and impact liver function.(25–27) A prospective study of LT patients demonstrated that the majority were colonized with multidrug-resistant organisms (MDROs), which was associated with increased rates of MDRO infections. (1) A follow-up study demonstrated that colonization with MDROs was associated with reduced microbiota diversity, as determined by 16S rRNA gene sequencing. (2) How microbiota-derived metabolites contribute to colonization resistance against MDROs and whether metabolite profiling can identify patients at high risk for MDRO infections remains open questions.

To determine the dynamic between the microbiota, microbiota-derived metabolites, MDRO expansion in the gut and invasive infection in liver transplant recipients, we performed a prospective surveillance study assessing fecal microbial composition and microbe-derived metabolite concentrations. We demonstrate that expansion of MDRO populations in the fecal microbiota correlates with increased risks of infection and that patients with low concentrations fecal SCFA, secondary bile acids and a subset of indole compounds are more likely to have *Enterococcus* expansion. Further, specific microbiome derived metabolite profiles identify patients with lost diversity as well as postoperative bacterial infection. Metabolomic profiling provides many advantages over metagenomic methods which are much slower and more expensive.

## RESULTS

### Microbiome compositions in liver transplant patients is widely variable

We conducted a prospective fecal microbiome and metabolome study on patients admitted to the University of Chicago Medical center for liver transplantation and enrolled 158 patients, of whom 28 did not undergo transplantation and 23 did not provide a fecal sample from 7 days prior to 30 days after transplantation (Supplementary Figure 1). For the 107 patients who underwent LT and from whom fecal samples were collected, the causes of end-stage liver disease were varied and included alcoholic cirrhosis/hepatitis (56%), malignancy (21%), non-alcoholic fatty liver disease (15%), and others (Table 1).

**Table 1.** Demographic and clinical characteristics of LT patients with and without infection. Variables represented as number and percent or median and interquartile range. Other indication for transplant includes: congestive hepatopathy, primary biliary cirrhosis, and unidentified. Immunosuppressive medications were included if given within the 30 day post operative period. Antibiotics were included if given in the first 7 pre-operative days or the day of LT. p-values obtained by Chi-squared goodness of fit and adjusted for multiple comparisons. Abbreviations: AI-American Indian; AN-Alaska Native; NAFLD-Non-Alcoholic Fatty Liver Disease; NASH-Non-Alcoholic Steatohepatitis; PSC-Primary Sclerosing Cholangitis; DILI-Drug Induced Liver Injury; MELD-Na – Model for End State Liver Disease Na; Pip/Tazo – Piperacillin/Tazobactam; IV-Intravenous; PO – *per orem*.

|  | No Infection | Infection | p |  | No Infection | Infection | p |
| --- | --- | --- | --- | --- | --- | --- | --- |
| **Demographics # (%)** |  |  |  | **Disease Severity # (%)** |  |  |  |
| Male | 44 (53.7) | 16 (64.0) | 0.88 | MELD-Na med [IQR] | 28 [19,33] | 30 [19,33] | 0.5 |
| Age med [IQR] | 55 [41,63] | 59 [46,68] | 0.35 | Dialysis | 22 (27.2) | 9 (34.6) | 1 |
| **Race** |  |  | .88 | Pressers | 6 (7.4) | 5 (19.2) | 1 |
| White | 54 (65.4) | 17 (65.4) |  | Ventilation | 5 (6.2) | 2 (7.7) | 1 |
| Black/African-American | 9 (1.1) | 2 (7.7) |  | **Immunosuppression** |  |  |  |
| More than one Race | 8 (9.9) | 2 (7.7) |  | Basiliximab | 54 (66) | 18 (72) | 1 |
| Asian/Mideast Indian | 6 (7.4) | 2 (7.7) |  | Tacrolimus | 82(100) | 25(100) | 1 |
| AI or AN | 1 (1.2) | 0 (0.0) |  | Mycophenolate | 72(89) | 24(96) | 1 |
| Declined | 4 (4.9) | 1 (3.8) |  | Corticosteroid | 82(100) | 25(100) | 1 |
| Unknown | 0 (0.0) | 2 (7.7) |  | **Antibiotics** |  |  |  |
| **Indication** |  |  |  | Pip/Tazo | 70 (85) | 23 (92) | 1 |
| Alcoholic Cirrhosis | 40 (48.8) | 8 (32) | 0.88 | Cefepime | 26 (32) | 11 (44) | 1 |
| Malignancy | 17 (21.0) | 6 (23.1) | 1 | Ceftriaxone | 16 (20) | 7 (28) | 1 |
| NAFLD/NASH | 10 (12.3) | 6 (23.1) | 0.88 | Meropenem | 4 (4.9) | 1 (4.0) | 1 |
| Alcoholic Hepatitis | 6 (7.4) | 2 (7.7) | 1 | Ciprofloxacin | 15 (18) | 4 (16) | 1 |
| PSC | 6 (7.4) | 0 (0.0) | 0.88 | Vancomycin IV | 42 (51) | 17 (68) | 1 |
| Autoimmune | 5 (6.2) | 0 (0.0) | 0.88 | Vancomycin PO | 1 (1.2) | 1 (4.0) | 1 |
| Cryptogenic | 4 (4.9) | 1 (3.8) | 1 | Metronidazole | 34 (42) | 12 (48) | 1 |
| Acute Viral Hepatitis | 4 (4.9) | 0 (0.0) | 0.88 | Rifaximin | 37 (45) | 12 (48) | 1 |
| Chronic Hepatitis C | 4 (4.9) | 0 (0.0) | 0.88 | Gentamicin | 1 (1.2) | 1 (4.0) | 1 |
| Wilson's Disease | 2 (2.5) | 1 (3.8) | 1 | Tobramycin | 4 (4.9) | 3 (12) | 1 |
| Chronic Hepatitis B | 0 (0.0) | 1 (3.8) | 0.88 | Daptomycin | 1 (1.2) | 3 (12) | 0.83 |
| Hemochromatosis | 0 (0.0) | 1 (3.8) | 0.88 | **Total** | 82 | 25 |  |
| DILI | 1 (1.2) | 0 (0.0) | 1 |  |  |  |  |
| Other | 8 (9.8) | 5 (20.0) | 0.88 |  |  |  |  |

We determined fecal microbiota compositions in LT recipients by shotgun metagenomic sequencing in the peri-transplant period and implemented MetaPhlAn-4 for determining the relative composition of microbial communities. (28) Microbiome composition and diversity varied widely between the 107 patients, with some approximating healthy donors (HD) and others being dominated by single bacterial taxa (Figure 1). To determine whether microbiome compositions influence post-transplant outcomes, we divided LT patients into three groups on the basis of microbiota diversity, with 27 high diversity patients falling within the range of diversities detected in the HD group and the remaining 80 LT patients were evenly divided into low and medium diversity groups. A taxUMAP of the 107 LT patients and 21 healthy donors revealed close clustering between the subset of high diversity LT and HD fecal samples while most of LT patient samples harbored microbiota with distinct and wide-ranging compositions (Figure 1). While high diversity LT patient and HD microbiome compositions shared some similarities, LT patients had significantly lower relative abundance of *Oscillospiraceae* (synonym *Ruminococcaceae*) (*W*(48) = 117, *p* < 0.001, two-tailed test), and higher abundance of *Enterobacterales* (*W*(48) = 487, *p* < 0.001, two-tailed). In contrast, low and medium diversity LT patients differed from high diversity patients in nearly every measured metric. In addition to reduced diversity, the low and medium diversity groups had lower abundances of the phylum Bacteroidetes (*W*(64) = 57, *p* < 0.001, two-tailed; *W*(67) = 342, *p* = 0.014, two-tailed, respectively) and families *Lachnospiraceae* (*W*(64) = 0, *p* < 0.001, two-tailed; *W*(67) = 140, *p* < 0.001, two-tailed, respectively) and *Oscillospiraceae* (*W*(64) = 32, *p* < 0.001, two-tailed; *W*(67) = 190, *p* < 0.001, two-tailed, respectively). Increases in *Enterococcus* and *Enterobacterales* (p<0.001) abundance were common in low and medium diversity LT patients. Many increases amounted to >90% of the entire composition of the microbiome, with 40% of LT patients having greater than 20% *Enterococcus* relative abundance and 17% of patients having greater than 5% *Enterobacterales* relative abundance. To ensure stool samples corresponded to the perioperative period, a histogram of stool sample collections was created (Supplemental Figure 2). The median day to stool collection was post-operative day 4, with most samples being collected within the first 7 days before or after transplant. There was no difference in stool collection timing between diversity groups.

**Figure 1.**
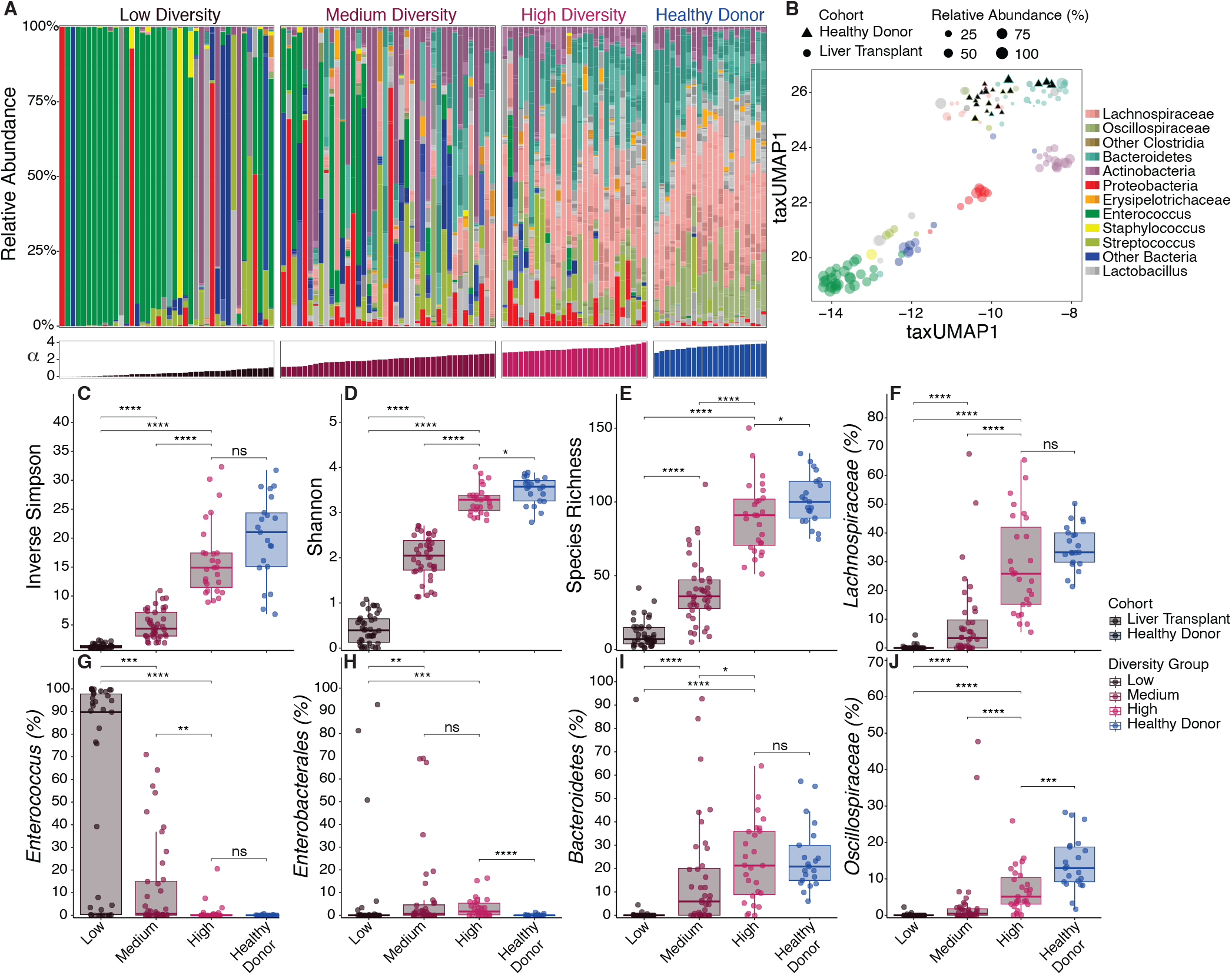
Microbiome compositions of LT recipients vary widely. (A) Fecal microbiome composition plots of liver transplant (LT) patients and healthy donors (HD) vertically organized by relative abundance and color coded by taxa. Individual samples were ordered horizontally by Shannon diversity. (B) taxUMAP1 plot of taxonomic composition, HD are denoted as triangles. Samples are color coded by the most abundant taxon and size determined by the relative abundance of that taxon. (C-E) Comparison of Alpha diversity between LT diversity groups and between high diversity and HD using (C) Inverse Simpson, (D) Shannon, and (E) Richness. (F-J) Comparison of relative abundance of select taxa between LT diversity groups and HD. (F) *Lachnospiraceae*; (G) *Enterococcus*; (H) *Enterobacterales*; (I) *Bacteroidetes*; (J) *Oscilospiraceae*. Significance tested by Kruskal-Wallis test *p≤ 0.05 **p≤0.01 ***p≤0.001 ****p≤0.0001

Although reduced abundance of beneficial taxa and expansion of *Enterococcus* and *Enterobacterales* species are known to occur in chronic liver disease and LT patients, the consequences of these compositional shifts on the microbiota-derived metabolome remain undefined. (1–4) We performed targeted GC- and LC-MS analyses for metabolites produced or modified by the gut microbiota on peri-transplant fecal samples from the LT patient cohort (Figure 2). Within the LT patient cohorts, relative amounts of butyrate, valerate, and hexanoate varied dramatically, with patients in the low and medium diversity cohorts having markedly reduced levels while those in the high diversity group approached the range seen in the healthy donors.

**Figure 2.**
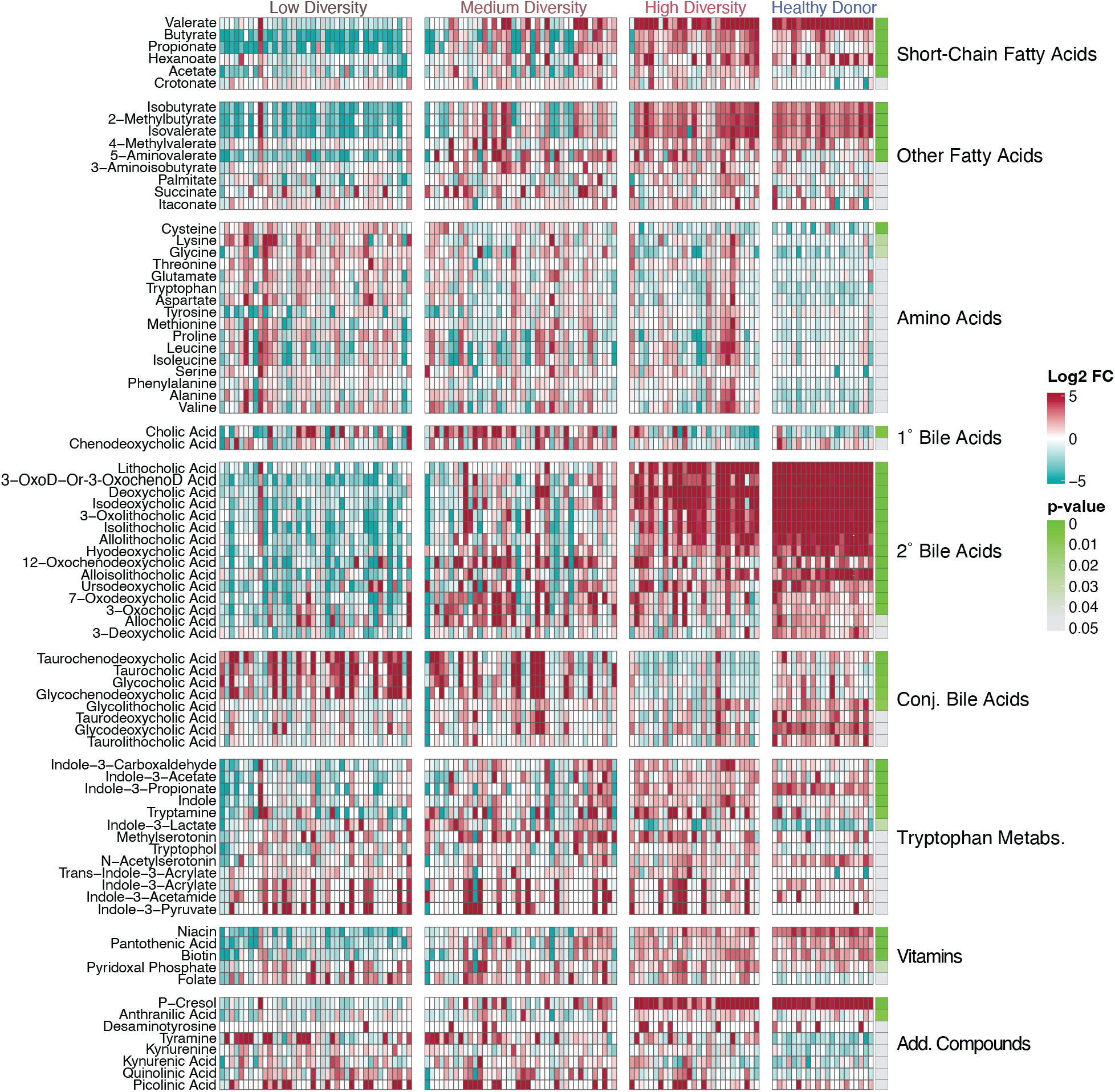
Qualitatively measured microbiome derived fecal metabolites vary widely among LT recipients. Individual metabolite abundances represented on a colorimetric heat map by log2 fold change from the mean between samples. Red indicating increased abundance; blue indicating reduced abundance. Significance was measured between LT groups using the Kurskal-Wallis test and denoted on a colorimetric scale where green represents lower p-values, adjusted for multiple comparisons. HD were included for visual comparison. Add. Compounds include kynurenine pathway and phenolic aromatics. Abbreviations: 1°-Primary; 2°-Secondary; Add.-Additional; 3-Oxo-D-Or-3-Oxocheno.-1-Oxo-Deoxycholic or 3-Oxochenodeoxycholic acid, which could not be completely discriminated chromatographically and are included together.

We detected markedly reduced concentrations of secondary bile acids and corresponding increases in concentrations of conjugated and primary bile acids among low and medium diversity LT patients, indicating loss of microbial bile acid deconjugating and 7*α*-dehydroxylation capacity. (29, 30) Fecal samples from LT recipients had increased abundance of amino acids, however this increase did not correlate with microbiota diversity or composition and thus may be a consequence of chronic liver disease and/or recent liver transplantation. Microbiota derived vitamins biotin, pantothenic acid, niacin, and folate were reduced in low diversity LT patients, likely reflecting loss of gut commensal bacteria that produce B vitamins, in particular species belonging to the phylum Bacteroidetes. (31) *P*-Cresol, a microbiome derived phenol which has been linked to cardiovascular risk in chronic kidney disease was also reduced in low diversity LT patients. (32) Only one microbiome derived metabolite, tyramine, which is produced by *Enterococcus faecalis* and *Enterococcus faecium*, is significantly more abundant in liver transplant recipients. (33)

Mass spectrometric quantitation of acetate, propionate and butyrate concentrations in fecal samples from LT patients revealed marked reductions in patients with low microbiota diversity [Figure 3] while the high diversity LT patients had SCFA concentrations in the same range as fecal samples from healthy subjects. Conjugated and unconjugated primary bile acids were present at very low concentrations in fecal samples from healthy subjects and high diversity LT patients while several low and medium diversity fecal samples had much higher concentrations. In contrast, fecal secondary bile acid concentrations were markedly lower in low and medium diversity compared to high diversity samples. Quantitation of fecal metabolites reveals the remarkable range of metabolic activity in the lower intestinal tract microbiota of LT patients, a the subset of patients with near normal microbiota diversity having metabolite concentrations within the normal range while the majority of LT patients, in particular those with the lowest microbiota diversities, have markedly reduced and in many cases absolute absence of beneficial metabolites.

**Figure 3.**
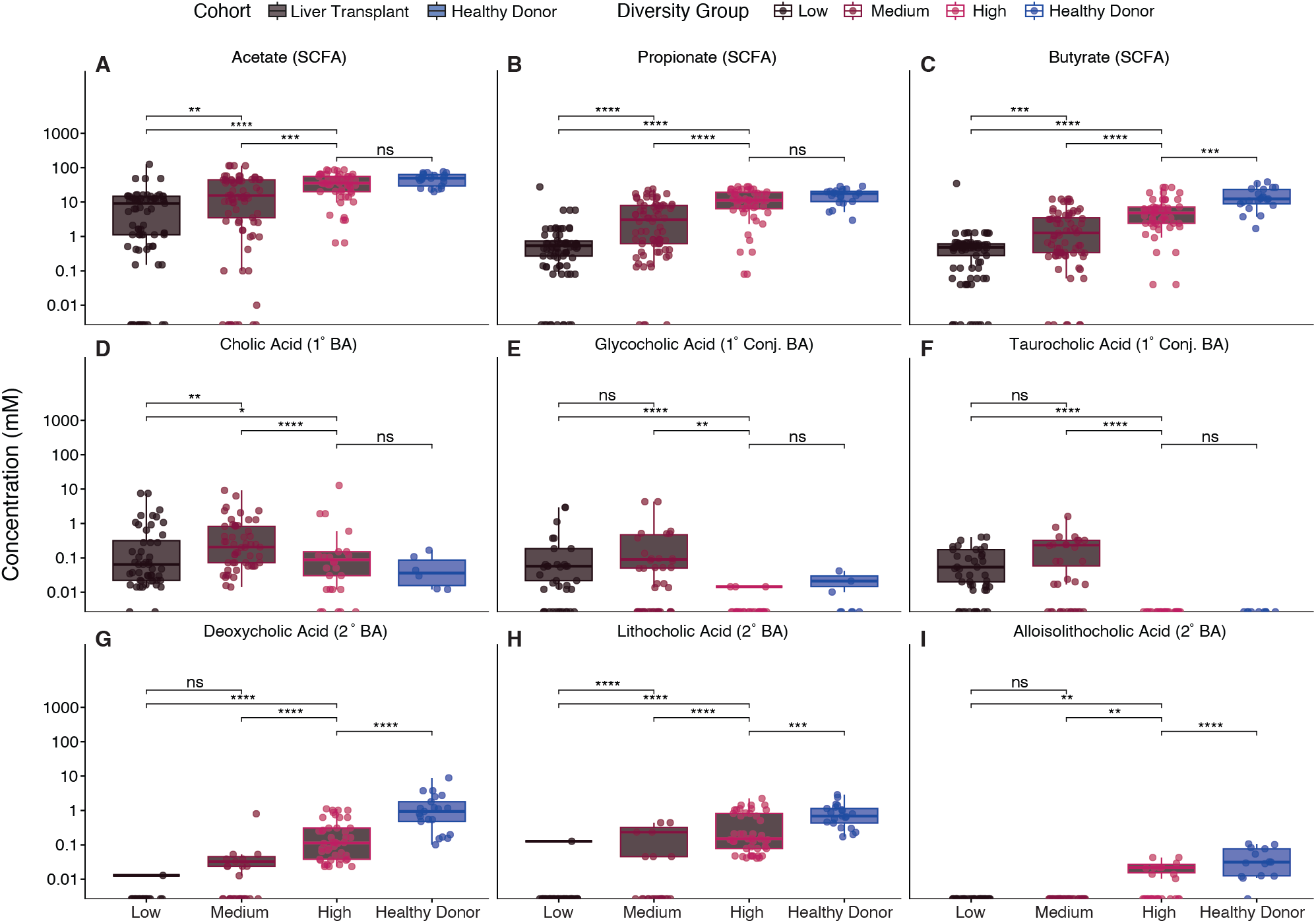
Quantitatively measured microbiome derived fecal metabolites vary widely among LT recipients. Absolute abundances of acetate, propionate, butyrate, cholic acid, glycocholic acid, taurocholic acid, deoxycholic acid, lithocholic acid, and alloisolithocholic acid compared between LT diversity groups and between high diversity LT and HD. *p≤ 0.05 **p≤0.01 ***p≤0.001 ****p≤0.0001

To determine whether taxonomic changes in microbiome composition or changes in microbial metabolism impact the risk for postoperative infection, we evaluated LT patients for infections in the first 30 days following transplant. Of the 107 LT recipients, 25 developed bacterial infection. [Supplemental Figure 1] We compared the taxonomic composition of stool samples collected closest to the time of infection and compared them to peri-transplant samples from patients who did not develop infection. There were no significant differences in the age, sex, or race between patients with or without infection. [Table 1] Additionally, there were no differences in indication for transplant, Model of End Stage Liver Disease-Na (MELD-Na) score, use of mechanical ventilation, renal replacement therapy, vasopressors, immunosuppression, or antibiotic administration between the groups. The most common site of infection was intra-abdominal (50%), followed by urinary tract (20%), and skin (13%). [Supplemental Table 1] The most common organisms causing infection were members of the genus *Enterococcus* (45%) and order *Enterobacterales* (20%). No specific organism was identified in 5 (13%) infections.

### Expansion of Enterococcus and Enterobacterales is associated with postoperative infection

Because *Enterococcus* and *Enterobacterales* were the most common causes of infection, we determined whether increases of these taxa was associated with the risk of invasive infection. Perioperative fecal samples were ordered by relative abundance of *Enterococcus* or *Enterobacterales* and compared to causes of infection. [Figure 4]. Infections caused by *Enterococcus* clustered with fecal samples with increased densities of *Enterococcus*, while *Enterobacterales* infections occurred in patients with increased fecal *Enterobacterales* density (Figure 4). However, many patients with expansion of pathobiont species did not develop invasive infections, suggesting that other unidentified factors, such as immune and mucosal barrier defenses and other clinical factors likely also contribute to the risk of infection.

**Figure 4.**
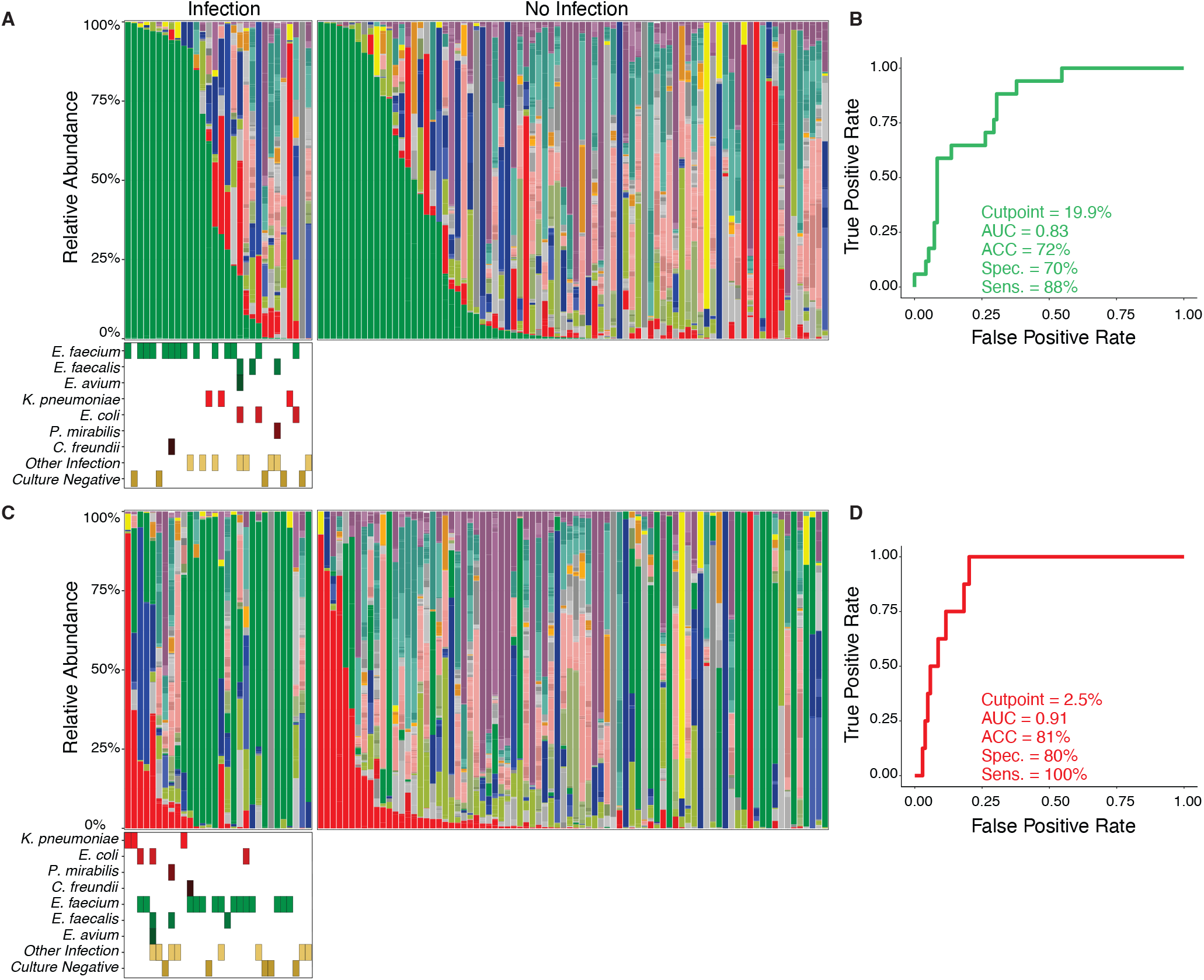
*Enterococcus* and *Enterobacterales* expansion in the gut microbiome predicts postoperative infection. (A) Fecal microbiome composition plots color coded by taxon. Plots are categorized by presence of bacterial infection and ordered by descending relative abundance of *Enterococcus*. Colored tiles indicate an infection caused by the denoted organism associated with that stool sample. For taxonomic color palate refer to Figure 1. (B) Receiver operator curve using Enterococcus abundance to predict *Enterococcus* infection. Cut point determined by Youden Index to optimize both sensitivity and specificity. 95% Confidence intervals for Accuracy (0.74-0.91), Specificity (59-94%), and Sensitivity (65-100%). (C) Fecal microbiome composition plots organized by relative abundance of *Enterobacterales*. (D) Receiver operator curve using *Enterobacterales* abundance to predict *Enterobacterales* infection. 95% Confidence intervals Accuracy (0.82-0.96), Specificity (72-94%), and Sensitivity (100-100%). Abbreviations: AUC-Area under the curve; ACC-Accuracy; Spec.-Specificity; Sens.-Sensitivity.

Receiver operator characteristic (ROC) analyses were performed to determine correlations between intestinal expansion of pathobionts with infection and optimized for sensitivity and specificity. The optimized threshold determined for *Enterococcus* relative abundances predicting *Enterococcus* infections was 19.9%. At this threshold expansion of *Enterococcus* predicts infection with sensitivity of 88% ([CI]: 65%-100%), specificity of 70% ([CI]: 59%-94%) and an area under the curve of 0.83 ([CI]: 0.74-0.91) (Figure 4). Expansion of *Enterococcus* >19.9% occurred in 41% of LT patients and accounts for 15 of the 17 (88%) of *Enterococcus* infections (Supplemental Table 2). For *Enterobacterales*, a threshold of 2.5%, relative abundance predicted infection with sensitivity of 100% ([CI]: 100%-100%), specificity of 80% ([CI]: 72%-94%) and an area under the curve of 0.91 ([CI]: 0.82-0.96) (Figure 4). *Enterobacterales* expansion at or above 2.5% occurred in 27% of LT patients and accounted for all 8 (100%) *Enterobacterales* infections.

### Microbiome derived metabolites associated with Enterococcus expansion

To determine whether fecal metabolite concentrations identify patients with *Enterococcus* or *Enterobacterales* expansion, we generated volcano plots comparing log2 fold changes of metabolites (relative to the median value of each compound) against statistical significance, with p-values adjusted for multiple comparisons following the Benjamini-Hochberg method (Figure 5). Expansion was defined as >=19.9% for *Enterococcus* and 2.5% for *Enterobacterales*, as determined in Figure 4. A variety of metabolites were enriched in samples without *Enterococcus* expansion and included several secondary bile acids: lithocholic, isolithocholic, allolithocholic, isodeoxycholic, alloisolithocholic, 3-oxolithocholic, and 12-oxolithocholic acids. Butyrate, valerate, propionate, and hexanoate were also enriched in patients without *Enterococcus* expansion. Other microbially derived metabolites that were enriched in patients without expansion included the tryptophan-derived metabolite indole-3-propionate, the vitamin biotin, and a number of branched chain fatty acids. Fifteen metabolites were enriched in patients with *Enterococcus* expansion, including multiple conjugated bile acids, likely indicating reduced microbiome-mediated bile acid deconjugation. Tyramine, a tyrosine derived amine produced by *E. faecium* and *E. faecalis* and several amino acids were also enriched in expanded samples. To confirm the trends established by qualitative analysis, a subset of metabolites with previously established beneficial effects were measured quantitatively against standardized controls. Of these, many of the same SCFA and secondary bile acids were enriched in samples without expansion. (Figure 5) The primary bile acid cholic acid and conjugated bile acid taurocholic acid were enriched in samples with expansion. The smaller number of LT patients with fecal *Enterobacterales* expansion precluded identification of statistically significant metabolite associations after correcting for multiple comparisons (data not shown).

**Figure 5.**
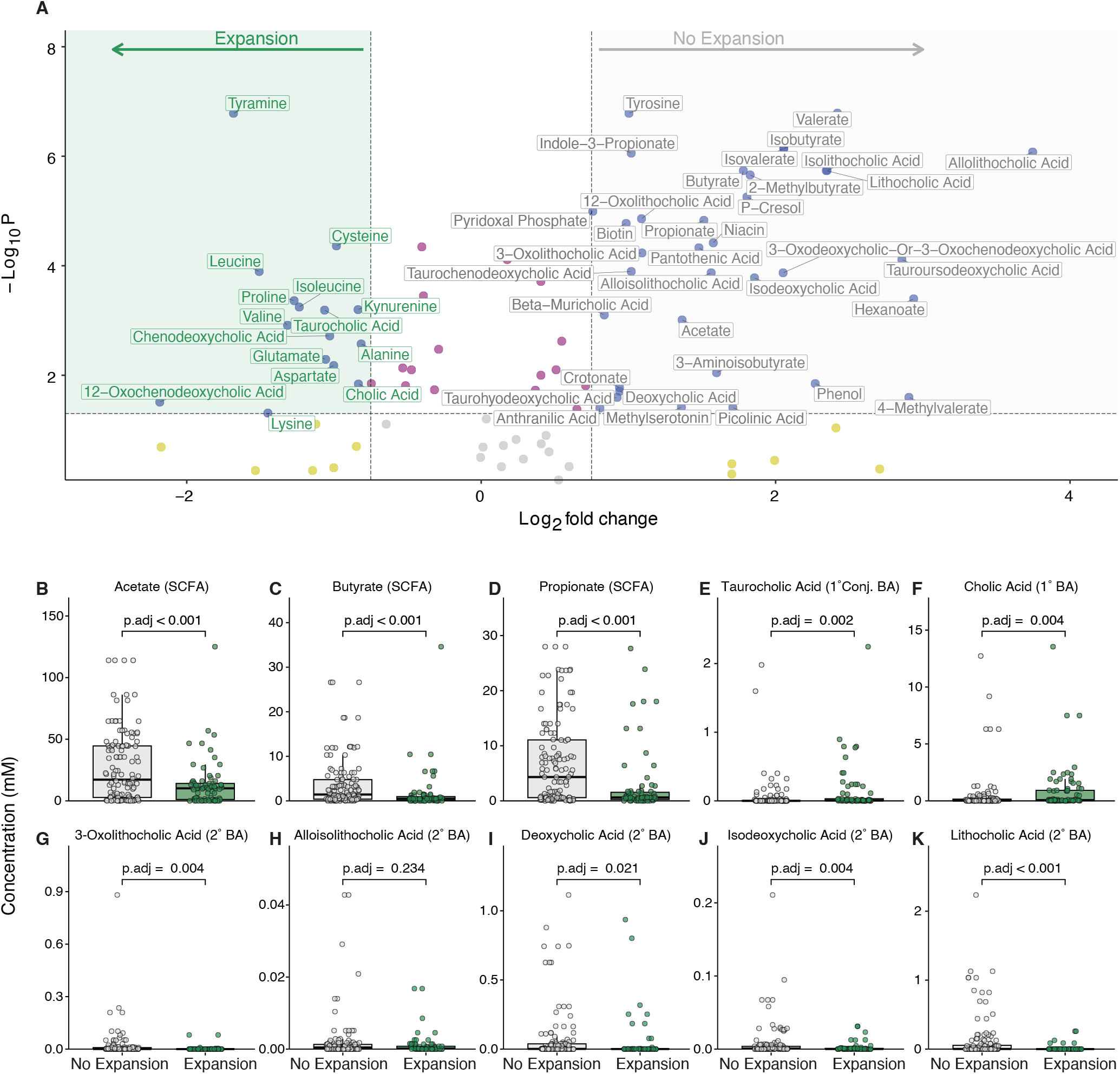
Microbiome derived fecal metabolites are enriched in LT recipients with and without *Enterococcus* expansion. (A) Volcano plot comparing Log2 fold change of qualitative metabolite concentration from the mean, and Log10 scale p-value adjusted for multiple comparisons. Only metabolites with Log2 fold change ≥+/-1 and p≤ 0.05 were labeled. (B-K) Quantitative metabolite comparison of select metabolites. (B) Acetate (C) Butyrate (D) Propionate (E) Taurocholic acid (F) Cholic acid (G) 3-Oxolithocholic acid (H) Alloisolithocholic acid (I) Deoxycholic acid (J) Isodeoxycholic acid (K) Lithocholic acid. Significance tested by Kruskal-Wallis test.

While microbiota composition and diversity correlate with fecal metabolomic profiles (Fig. 2), the ability of metabolomic profiling to predict microbiota compositions remains largely untested. Given the speed of metabolomic profiling compared to metagenomic sequence analysis and the interest in real-time identification of patients for potential microbiota reconstitution therapies, we decided to determine how accurately fecal metabolite concentrations predict microbiota diversity and the risk of infection. Using 93 metabolite measurements per fecal sample, sparse partial least squares discriminant analysis (sPLS-DA) (34) predicted low, medium, and high diversity groups with high sensitivity, specificity, and accuracy (Figure 6A) 70% ([CI]: 59.7%-78.3%). Secondary bile acids, short- and branched-chain fatty acids correlated most significantly with high diversity microbiota (Supplementary Figure 3). While the model most accurately identified low diversity fecal samples, only misidentifying 1 of 37 as high diversity, 7 of 26 high diversity samples were identified as low diversity, suggesting that exogenous factors such as diet may influence the metabolic activity of resident commensal bacteria.

**Figure 6.**
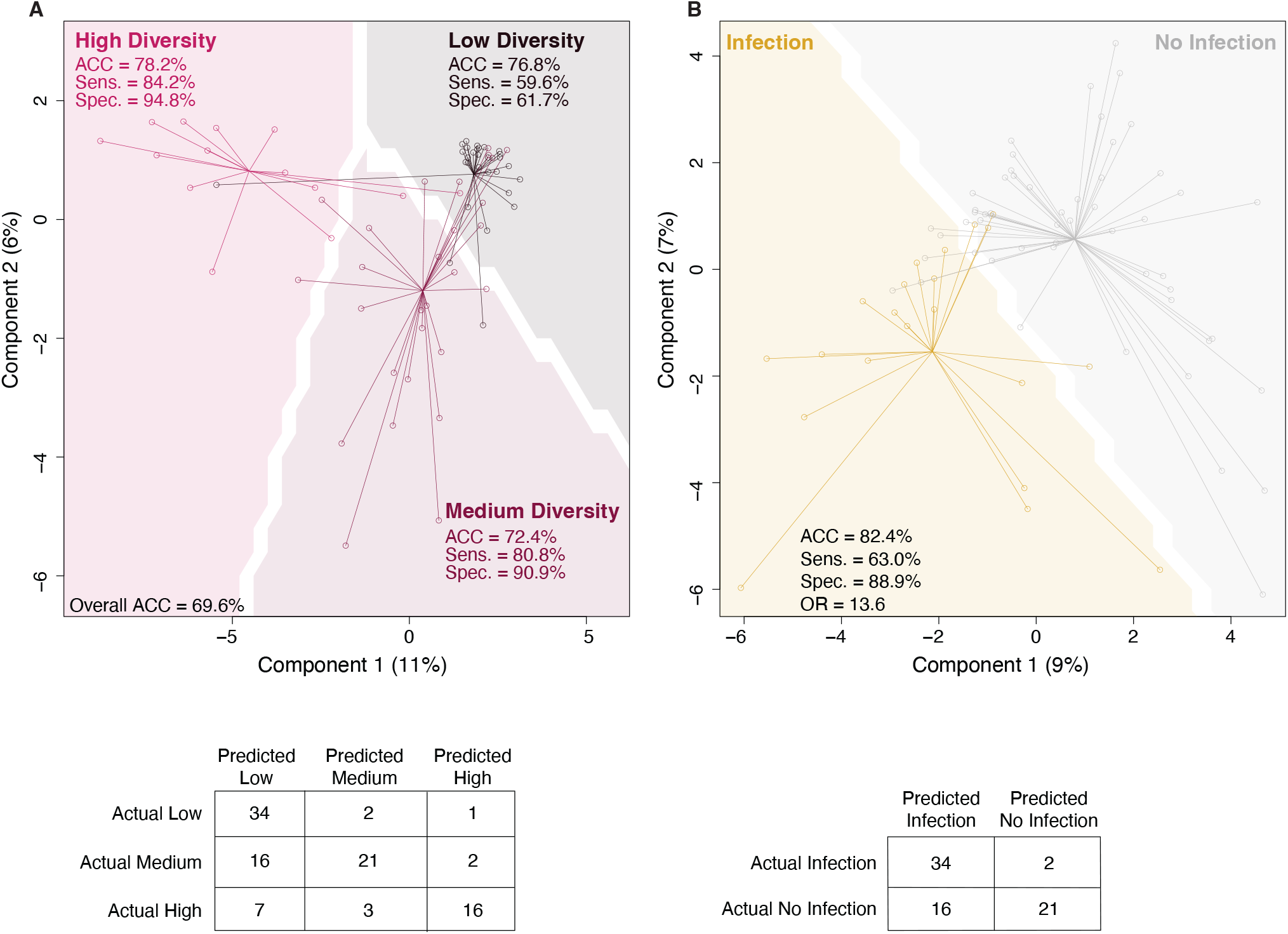
Microbiome derived fecal metabolites identify low, medium, and high diversity samples and post operative infection. (A) sPLS-DA using input matrix of sample metabolites and predicted microbial diversity group. Comparison between predicted groups was visualized on a grid with dividing lines and optimized by maximum distance between groups. Accuracy was: Low Diversity 77%, Medium Diversity 72%, and High Diversity 78%. Sensitivity ranged from 60-84%. Specificity ranged from 62-95%. (B) sPLS-DA using input matrix of sample metabolites and predicted postoperative infection. Comparison between outcomes was visualized on a grid with a dividing line and optimized by maximum distance between groups. Accuracy was 82.2% [73.9-89.1%], sensitivity was 63% [42.4-80.6], specificity was 88.9% [80-94.8%], and odds ratio was 13.6 [4.8-38.6].

Fecal metabolite levels also accurately predict pathobiont dissemination beyond the gut to sites of postoperative infection such as the peritoneal cavity, urinary tract, or bloodstream (Figure 6B). Samples obtained from patients with infection clustered distinctly from those without infection, with an overall accuracy of 82% ([CI]: 73.9%-89.1%), a strong odds ratio (13.6 ([CI]: 4.8-38.6) and high specificity of 89% ([CI]: 80%-94.8%). Sensitivity for infection was slightly lower 63% ([CI]: 42.4%-80.6%), likely reflecting multifactorial causality of infection following LT. To determine which metabolites made the greatest contribution to the PLS-DA models, metabolite loadings for each axis were assembled in separate plots (Supplemental Figure 4). The largest contributors to the model’s first two components were tyramine, conjugated bile acids, indole derivatives, and SCFAs in patients who remained uninfected.

## DISCUSSION

Every LT patient follows a distinct path from health to end-stage liver disease (ESLD). Some patients develop acute, fulminant hepatitis over the course of days, while others progressively lose hepatic function over many years due to chronic, progressive fibrosis leading to cirrhosis and end stage complications. The wide range of microbiome compositions we and others have detected in LT cohorts (1–4) likely result from diversity of diseases leading to ESLD, differing treatments modalities, including exposure to antibiotics for infections such as spontaneous bacterial peritonitis and acquisition of antibiotic-resistant pathobionts. We demonstrate that targeted mass spectrometric analysis of fecal samples identify LT patients with partial to complete loss of metabolites that impact pathobiont fitness, host immune defenses, and mucosal barrier integrity. Loss of beneficial metabolites closely correlates with loss of bacterial taxa that constitute a diverse microbiota and identifies patients with an increased risk of postoperative infections caused by antibiotic-resistant pathobionts.

The ability of the intestinal microbiota to protect against enteric infection has been appreciated for over 70 years, with Bohnhoff et al. and Freter et al. (35, 36)demonstrating that antibiotic treatment alters the intestinal microbiota and renders mice markedly more susceptible to infection by enteric pathogens. Subsequent experimental studies demonstrated that SCFA and secondary bile acid production by obligate anaerobic commensal bacterial species suppress enteric pathogens and pathobionts. (22–24, 36) Microbiota also enhance immune defenses in the gut through induction of IL-23 secretion by lamina propria dendritic cells, which drives IL-22 production by innate lymphocytes which in turn stimulates growth of epithelial stem cells and production of antibacterial C-type lectins such as RegIII*γ*. (37)These lectins inhibit many human pathogens, including *Enterococcus faecium, Escherichia coli*, and other enteric gram negatives. In parallel, production of indole compounds by microbiota-mediated tryptophan metabolism drives differentiation of immune cells by triggering the aryl-hydrocarbon receptor (AHR) and enhancing IL-22 production. (38) Our findings support the notion that a diverse microbiome, with the capacity to produce SCFAs, secondary bile acids and AHR ligands, enhances resistance of LT patients to pathobiont expansion in the gut lumen and invasive infection from the gut.

The contributions of commensal bacterial species and their metabolic products to disease resistance in clinical settings are challenging to disentangle. Treatment with broad-spectrum antibiotics, the most common cause of microbiome compromise, was very common in our cohort, which results in concurrent depletion of many microbial taxa and microbiota-derived metabolites. (16, 39, 40) Loss of beneficial Bacteroidetes, *Lachnospiraceae* and *Oscillospiraceae* are paralleled by loss of SCFAs, secondary bile acids and indole compounds, making it impossible to pinpoint a specific deficiency that renders patients more susceptible to infection. It is likely that disease resistance is not fully attributable to any one component but instead results from the aggregate contributions of different components. While the specific mechanisms by which a diverse microbiota and resulting metabolites mediate protection from infection remain to be defined, the correlative results obtained from non-interventional studies suggest that clinical trials of therapeutic interventions to re-establish missing commensal bacterial populations and normalize metabolite profiles are warranted.

Metagenomic analyses of fecal samples require deep nucleic sequencing platforms and bioinformatic analyses that continue to evolve as bacterial genome databases grow, microbial nomenclature changes, and microbial gene annotation improves. Targeted metabolomic analyses by mass spectrometry, on the other hand, are rapid, accurate, and bioinformatic platforms to define features and provide chemical structures are well established. Our findings demonstrate that patients with compromised microbiomes, as defined by metagenomic analyses, can be identified by measuring fecal SCFAs, bile acids, and a narrow range of microbially generated metabolites. Not only can patients with reduced diversity be identified, but also those at increased risk for invasive bacterial infection. Given the rapidity with which microbiome compositions can change from one day to another, (41) obtaining same-day results would provide critical, real-time insights to guide clinical interventions to improve microbiome compositions and functions, such as therapy with live biotherapeutic products and/or microbiome modulating foods.

Our study has several limitations. Most importantly, while we demonstrate that the incidence of infection is associated with fecal microbiome compositions and metabolite concentrations, we are unable to prove causation. Future studies that intervene on microbiome and/or metabolite deficiencies are required to determine whether the reintroduction of specific bacterial taxa and/or their metabolic products reduces infection and resultant morbidity, mortality, and healthcare utilization. Our study is limited to one institution and, in contrast to other studies of microbiome compositions in LT patients, has a higher proportion of patients with ethanol associated liver disease and a low proportion of patients with hepatitis C related liver disease. While this makes our study distinct from prior studies, it closely represents cohort of contemporary patients who are being treated with transplantation. (42) The care of postoperative LT patients is very protocolized and center specific. Perioperative antibiotic use, nutrition, indications for transplant, hospital antibiogram, infection control practices, and many other factors are likely specific to our institution and may have influenced our findings. Thus, our findings may not directly extend to substantially different patient populations.

In conclusion, our study demonstrates that targeted fecal metabolite measurements can identify a large subset of patients undergoing LT with markedly deficient microbiome compositions. The ability to rapidly identify patients with an increased risk of postoperative infection should facilitate the development of strategies that preserve or reconstitute microbiome functions and lead to approaches that prevent exposure to antibiotic-resistant pathobionts. As administration of well-characterized microbes to reconstitute the microbiome evolves, rapid fecal metabolite monitoring will facilitate the development of personalized microbial consortia that correct metabolite deficiencies.

## METHODS

### Patient Enrollment

Patients eligible for LT or having recently undergone LT at University of Chicago Medicine were recruited to the protocol (IRB20-0163. Healthy subjects were recruited from the University of Chicago campus and were screened for absence of recent antibiotic treatment and autoimmune or other chronic illnesses prior to fecal microbiome and metabolome characterization.

### Definition of Postoperative Infection

All patient records were screened for documentation of clinical infection during the study period. Screening was performed by author CL at 1-month intervals. Infection was defined as any positive test for infection (microbiological, molecular, biochemical, or radiographic) that was documented as a true positive by the treating clinical team in the medical record. The majority of infections were microbiologically defined by culture or polymerase chain reaction. Infections were classified by location, microbiology, and antimicrobial resistance patterns (Supplemental Table 1). Infections were included for comparison if the infection occurred within the first 30 postoperative days and a stool sample was collected within 14 days prior or 2 days following the development of the infection. Each infection was paired with a stool sample collected nearest to the diagnosed infection. In patients who did not develop infection, the stool sample closest to the day of liver transplant was used for comparison.

### Inclusion/Exclusion

Patients were included in this analysis if they received a LT and had a stool sample collected within peri-operative period: days -7 to +30. Patients were excluded from analysis if they developed an infection, but a stool sample was not collected within 14 days preceding or 2 days following the diagnosis of the infection. 158 patients were enrolled in the study. 28 patients had not received a LT, 23 patients did not have a stool sample collected within postoperative day -7 to +30, and all patients had a stool sample collected within -14 to +2 days relative to the diagnosis of an infection (Supplemental Figure 1). The remaining 107 patients were included for analysis. 25 patients developed a bacterial infection and 82 did not. 40 unique infections occurred in the cohort because some patients developed more than one infection.

### Specimen Collection

After enrollment, stool samples were collected before and after LT. All stool samples were collected while admitted at University of Chicago Medical Center. Collection was started immediately after study enrollment and continued for up to two years. Each sample was immediately refrigerated and frozen within 24 hours of collection. Microbiota compositions and metabolite profiles were then characterized.

### Specimen Analysis

#### DNA Extraction

DNA was extracted using the QIAamp PowerFecal Pro DNA kit (Qiagen). Prior to extraction, samples were homogenized by mechanical disruption using a bead beater. Briefly, samples were suspended in a bead tube (supplied by Qiagen) with lysis buffer and homogenized on a bead mill (Fisherbrand). Samples were then centrifuged, and supernatant was resuspended in a reagent that effectively removed inhibitors. DNA was then purified routinely using a spin column filter membrane and quantified using Qubit 2.0 fluorometer.

#### Shotgun Sequencing

Libraries were prepared using 100-200 ng of genomic DNA using the QIAseq FX DNA library kit (Qiagen). Briefly, DNA was fragmented enzymatically into smaller fragments and desired insert size was achieved by adjusting fragmentation conditions. Fragmented DNA was end repaired and ‘A’s’ were added to the 3’ends to stage inserts for ligation. During ligation step, Illumina compatible Unique Dual Index (UDI) adapters were added to the inserts and the prepared library was amplified. Libraries were then cleaned up, and library size was measured using a TapeStation (Agilent). Sequencing was performed on the Illumina NextSeq 500 platform producing 5-10 million reads. Adapters were trimmed off from the raw reads, and their quality were assessed and controlled using Trimmomatic (v.0.39), (43) then human genome was removed by kneaddata (v0.7.10, https://github.com/biobakery/kneaddata). Taxonomy was profiled using metaphlan4 using the resultant high-quality reads. (35) Alpha diversity of fecal samples was calculated using the Inverse Simpson and Shannon indices. Taxonomic uniform manifold approximate projections (taxUMAP) were used to visualize beta diversity (https://github.com/jsevo/taxumap).

#### Metabolomic analysis

SCFAs were derivatized with pentafluorobenzyl bromide (PFBBr) and relatively quantified via negative ion collision induced-gas chromatography-mass spectrometry. Four SCFA’s were quantified absolutely: acetate, butyrate, succinate, and propionate ([-]CI-GC-MS, Agilent 8890). (44) Eight bile acids (primary: cholic acid; conjugated primary: glycocholic acid, taurocholic acid; secondary: deoxycholic acid, lithocholic acid, isodeoxycholic acid, alloisolithocholic acid, and 3-oxolithocholic acid) were quantified (μg/mL) by negative mode liquid chromatography-electrospray ionization-quadrupole time-of-flight-MS ([-]LC-ESI-QTOF-MS, Agilent 6546). Eleven indole metabolites were quantified by UPLC-QqQ LC-MS. Seventy additional compounds were relatively quantified using normalized peak areas relative to internal standards.

### Statistical Analysis

All analysis was conducted using R statistical language (v4.2.2 (2022-10-31)) using the RStudio (v2022.12.0+353) integrated development environment. Non-parametric, descriptive statistics, were used to describe the clinical characteristics of patients. All clinical tables were generated using the R package gtsummary. (45) For categorical clinical variables, a Chi-square goodness-of-fit test was implemented. All other continuous variables regressed against categorical variables (e.g. relative abundance vs diversity groups or age vs infection group) were statistically analyzed using a two-tailed, Wilcoxon-rank sum test, from the R package rstatix, (46) unless otherwise noted. All *p*-values were adjusted to account for multiple comparisons following the Benjamini-Hochberg method. Adjusted *p*-values of the tests were considered to be statistically significant for all analyses conducted if *p* ≤ 0.05. To compare relatively quantified metabolites, log2 fold-change (log2FC) was calculated per compound using the median value across all fecal samples. These log2FC values were then arranged by statistical significance when comparing the diversity groups (Kruskal-wallis test) and visualized on a heatmap using the ComplexHeatmap package (https://github.com/jokergoo/ComplexHeatmap). Volcano plots, visualized via the R package EnhancedVolcano (https://github.com/kevinblighe/EnhancedVolcano), used the same log2FC method as mentioned, but were statistically compared using two-tailed, Wilcoxon-rank sum tests. Receiver operator characteristic curves (ROC) were generated using the R package cutpointr (47) with the predicting variable as relative abundance of either *Enterococcus* or *Enterobacterales* and the outcome variable as *Entercooccus* or *Enterobacterales* infection, respectively. ROC curves were optimized for the Youden index and visualized using ggplot2. (48) Sparse partial least squares discriminatory analysis (sPLS-DA) was constructed using the R package mixOmics and required an input matrix of samples by metabolites. Models were tuned using 5-fold cross-validation repeated 50 times (mixOmics::perf and mixOmics::tune.splsda). The main hyperparameters that required tuning were the number of optimal components to retain in the model, number of metabolites to keep in the final model, as well as distance metric, for which we used the maximum distance “ max.dist”. The final model was built (mixOmics::splsda) and was used to predict classes of either diversity group or infection (predict). Background predictions (shaded areas on Figure 6), were constructed using the final sPLS-DA model (mixOmics:: background.predict). Additional model metrics for predicting bacterial infection were obtained via the R package caret::confusionMatrix (49) or epiR::epi.tests. (50) Since the model for predicting diversity groups included a multi-class prediction, additional model metrics were obtained from using the R package mltest::ml_test. (51) The inclusion flow diagram was built using the R package PRISMAstatement::flow_exclusions. (52) All data and code are available on both Github (https://github.com/DFI-Bioinformatics/Microbiome_Liver_Transplant) and Zenodo (https://zenodo.org/badge/latestdoi/592530758).

## Data Availability

All data and code are available on both Github (https://github.com/DFI-Bioinformatics/Microbiome_Liver_Transplant) and Zenodo (https://zenodo.org/badge/latestdoi/592530758). Shotgun metagenomic sequences can be found on NCBI via this accession number: PRJNA928758.

https://zenodo.org/badge/latestdoi/592530758

https://www.ncbi.nlm.nih.gov/bioproject/PRJNA928758

## Study approval

This study received approval by the institutional review board at the University of Chicago for human subjects research (IRB20-0163). At the time of enrollment, all study procedures were explained and written informed consent was obtained from the subject and/or an approved surrogate decision maker. Healthy donors were identified separately from LT patients. Written informed consent was also obtained from HD prior to participation.

## AUTHOR CONTRIBUTIONS

CJL designed research studies, obtained ethical approval, enrolled patients, developed sample collection and handling protocols, analyzed data, edited figures, wrote the manuscript. NPD performed, designed, and modified all data analysis, built figures, performed statistical analysis, and influenced study direction. MO enrolled patients, collected samples, and contributed to study direction. RN built patient enrollment, sample collection, and sample processing protocols. MK enrolled patients, collected samples, and abstracted data. JB enrolled patients, collected samples, and abstracted data. EA enrolled patients, collected samples, abstracted data, and maintained ethical approval. MRS contributed to study direction. MDC contributed to study direction. AM contributed to study direction. HL contributed to study direction, analyzed data, and edited figures. AS performed metagenomic sequencing of all samples. JLC assisted in metabolomic measurements. AMS performed metabolomic measurements. EGP designed research studies, provided guidance of patient population, clinical questions, sample collection protocols, metagenomic sequencing methods, metabolomic profile design, and secured funding for all studies. AA influenced study design, and guided clinical associations. JF influenced study design and guided clinical associations. TBB initiated the study in LT patients, identified clinical end points, helped establish recruitment and sample collection. AK designed research studies, established clinical end points, maintained recruitment and sample collection, and guided study findings.

## ACKNOWLEDGMENTS

Thank you to the Duchossois Family Institute, for establishing the metagenomic and metabolomic platforms used in this study, funding the study, and guiding the mission of the research.

---

**Supplemental Figure 1.**
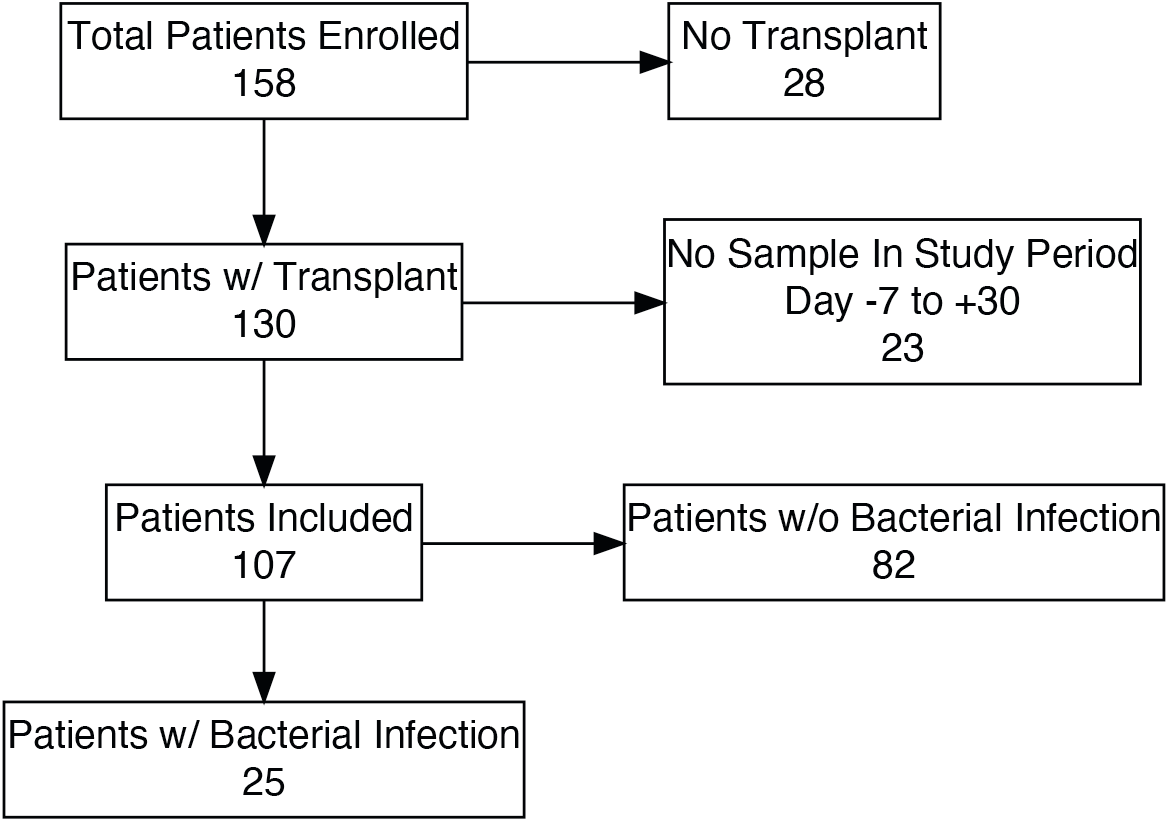
Flow diagram of inclusion and exclusion criteria. 28 patients were excluded because they had not yet undergone a LT. 23 patients were excluded because a sample wasn’t collected within -7 to 30 days post LT. 107 patients were included for analysis, 25 developed postoperative bacterial infection, 82 did not.

**Supplemental Figure 2.**
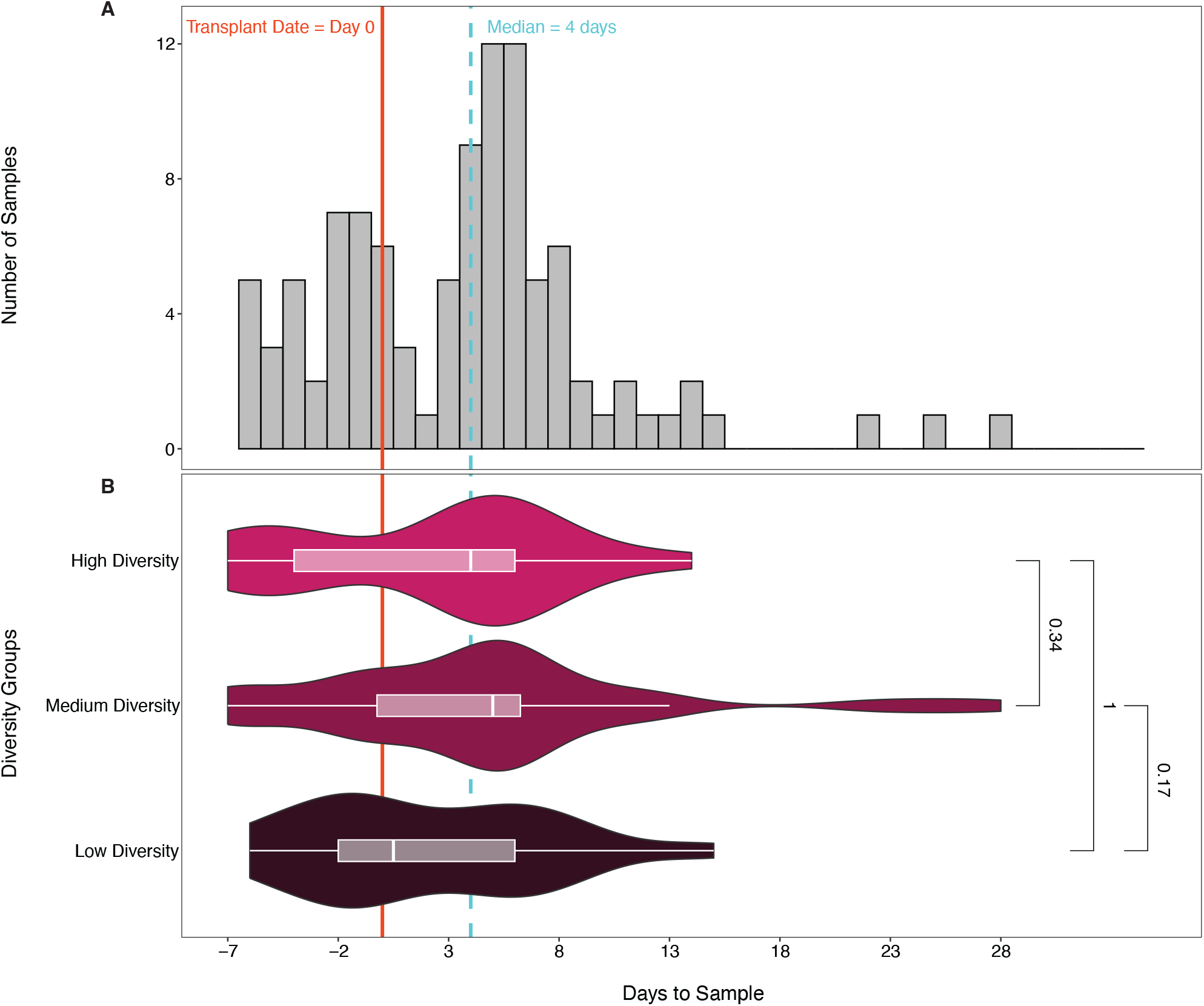
Time to first stool collection relative to day of liver transplant. (A) Histogram of first stool collections normalized to the day of transplant. The median being post operative day 4. The majority falling between day -7 and +7. (B) Median and IQR of day to first stool sample collected by diversity group. No significant difference existed between groups, suggesting perioperative antibiotics don’t explain the difference in microbiome composition between groups.

**Supplemental Table 1.** Characteristics of infections. *Enterococci* accounted for most infection, 45%. Followed by *Enterobacterales* 20%. The remaining 35% of infection were caused by various other bacteria. The total number of organisms identified is 40, which is greater than the number of infections (30) because of polymicrobial infections. Abbreviations: Surg-Surgical.

|  | #(%) |
| --- | --- |
| ***Enterococcus*** | 18 (45) |
| *E. faecium* | 14 (35) |
| *E. faecalis* | 3 (7.5) |
| *E. avium* | 1 (2.5) |
| ***Enterobacterales*** | 8 (20) |
| *E. coli* | 3 (7.5) |
| *K. pneumoniae* | 3 (7.5) |
| *P. mirabilis* | 1 (2.5) |
| *C. freundii* | 1 (2.5) |
| **Other Bacteria** | 14 (35) |
| *S. aureus* | 3 (7.5) |
| *P. aeruginosa* | 2 (5) |
| *S. maltophilia* | 1 (2.5) |
| *Bacteroides sp.* | 1 (2.5) |
| *C. difficile* | 1 (2.5) |
| *H. pylori* | 1 (2.5) |
| Culture Negative | 5 (12.5) |
| **Site of Infection** |  |
| Abdominal | 15 (50) |
| Skin/Surg. Site | 5 (16.7) |
| Urinary | 6 (20) |
| Blood Stream | 2 (6.7) |
| Lower Airway | 2 (6.7) |

**Supplemental Table 2.** *Enterococcus* and *Enterobacterales* expansion in the gut microbiome predicts postoperative infection. (A) 44/112 (41%) of included patients had *Enterococcus* expansion **≥**19.9% or higher. 88% (15/17) of infectious caused by *Enterococcus* were associated with expansion of 19.9% or greater. Odds ratio for *Enterococcus* infection was 17.1. (B) 29/112 (27%) of included patients had *Enterobacterales* expansion of 2.5% or higher and all 100% (8/8) of infectious caused by *Enterobacterales* were associated with expansion of 2.5% or more. The Odds ratio for *Enterobacterales* approached infinity, as no infections occurred in those <2.5%. In combination, only 2/25 (8%) of *Enterococcus* or *Enterobacterales* infections occurred without any expansion.

| **A *Enterococcus*** | **≥19.9% Rel. Abd.** | **<19.9% Rel. Abd.** | **Total** |
| --- | --- | --- | --- |
| Infected | 15 | 2 | 17 |
| Uninfected | 29 | 66 | 95 |
| Sensitivity | 88.2% |  |  |
| Specificity | 69.5% |  |  |
| Odds Ratio | 17.1 |  |  |

| **B *Enterobacterales*** | **≥2.5% Rel. Abd.** | **<2.5 Rel. Abd.** | **Total** |
| --- | --- | --- | --- |
| Infected | 8 | 0 | 8 |
| Uninfected | 21 | 83 | 104 |
| Sensitivity | 100% |  |  |
| Specificity | 79.8% |  |  |
| Odds Ratio | Inf |  |  |

**Supplemental Figure 3.**
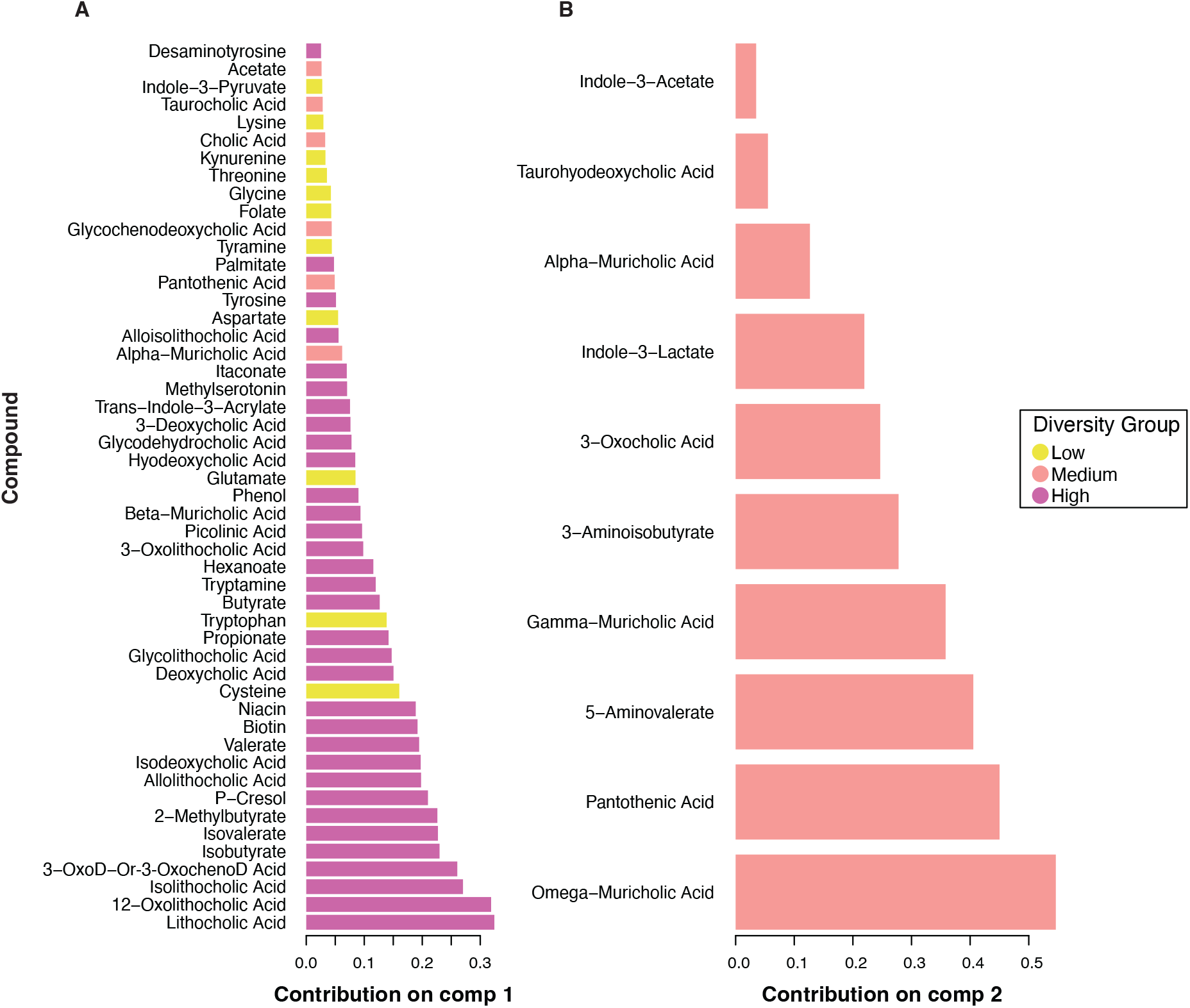
Microbiome derived fecal metabolites identify low, medium, and high diversity samples. Metabolite component loadings of sPLS-DA model (Figure 6A). x-axis indicates magnitude of impact on each component. Each component is color coded for which outcome it correlates, Low, Medium, and High diversity. 3-Oxodeoxycholic acid and 3-Oxochenodeoxycholic acid could not be discredited chromatographically, so are included together.

**Supplemental Figure 4.**
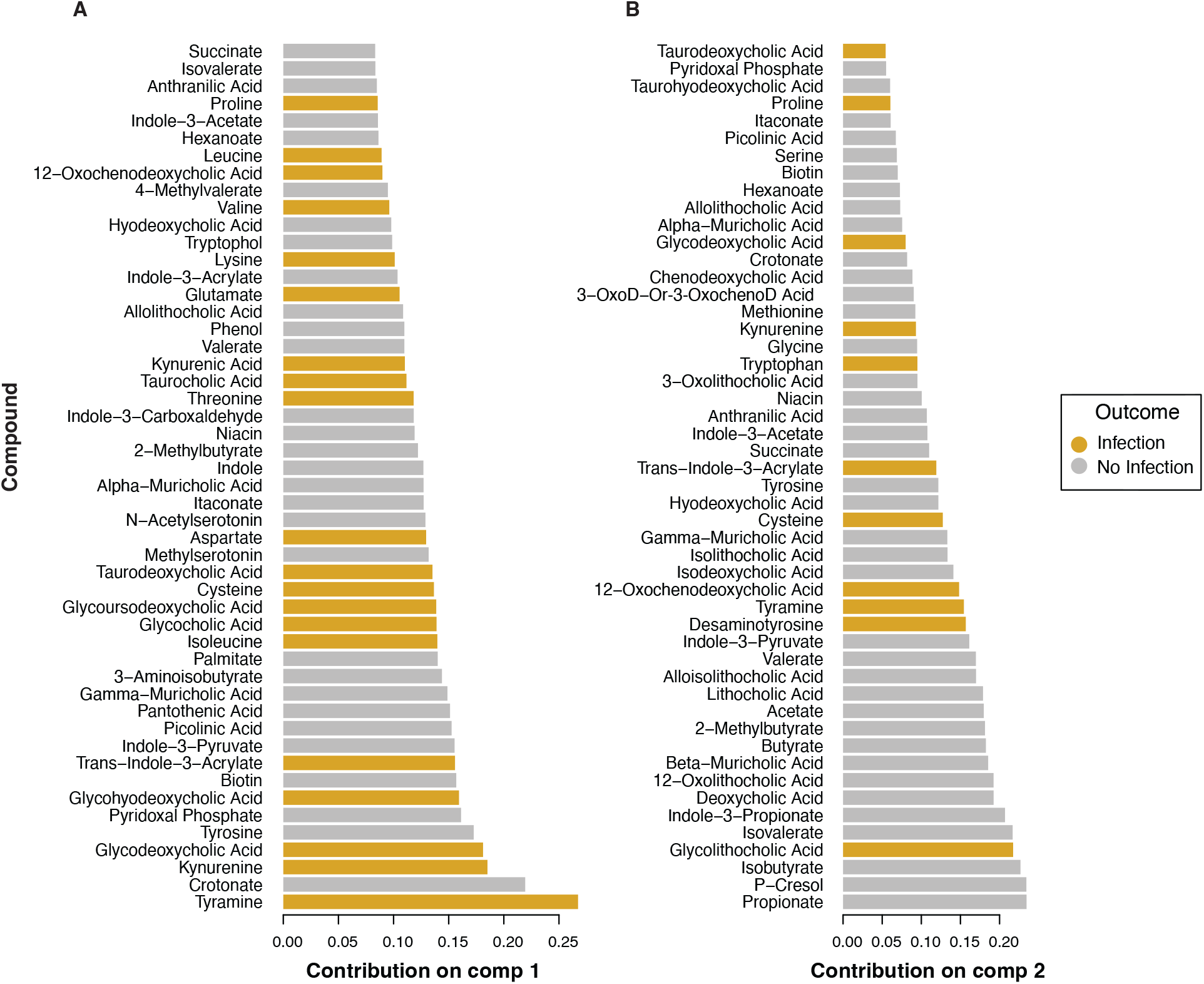
Microbiome derived fecal metabolites identify postoperative infection. Metabolite component loadings of first two components of sPSL-DA model predicting postoperative infection. (Figure 6B) The x-axis corresponds to the magnitude of impact each metabolite has on the component. Each metabolite is color coded for infection or no infection. 3-Oxodeoxycholic acid and 3-Oxochenodeoxycholic acid could not be discredited chromatographically, so are included together.

